# Morphometric Analysis of Spina Bifida after Fetal Repair Shows New Subtypes with Associated Outcomes

**DOI:** 10.1101/2024.05.29.24308088

**Authors:** Lovepreet K. Mann, Shreya Pandiri, Neha Agarwal, Hope Northrup, Kit Sing Au, Elin Grundberg, Eric P. Bergh, Mary T. Austin, Rajan Patel, Brandon Miller, Sen Zhu, Jonathan S. Feinberg, Dejian Lai, KuoJen Tsao, Stephen A. Fletcher, Ramesha Papanna

## Abstract

**Importance:** The binary classification of spina bifida lesions as myelomeningocele (with sac) or myeloschisis (without sac) belies a spectrum of morphologies, which have not been correlated to clinical characteristics and outcomes.

**Objective:** To characterize spina bifida lesion types and correlate them with preoperative presentation and postoperative outcomes.

**Design:** Secondary analysis of images and videos obtained during fetoscopic spina bifida repair surgery from 2020-2023.

**Setting:** Fetal surgery was performed at a quaternary care center.

**Participants:** A prospective cohort of patients referred for fetal spina bifida underwent fetoscopic repair under an FDA-approved protocol. Of 60 lesions repaired, 57 had available images and were included in the analysis.

**Intervention(s) or Exposure(s):** We evaluated lesion morphology on high-resolution intraoperative images and videos to categorize lesions based on placode exposure and nerve root stretching.

**Main Outcome(s) and Measure(s):** The reproducibility of the lesion classification was assessed via Kappa interrater agreement. Preoperative characteristics analyzed include ventricle size, tonsillar herniation level, lower extremities movement, and lesion dimensions. Outcomes included surgical time, need for patch for skin closure, gestational age at delivery, preterm premature rupture of membranes (PPROM), and neonatal cerebrospinal fluid (CSF) diversion.

**Results:** We distinguished five lesion types that differ across a range of sac sizes, nerve root stretching, and placode exposure, with 93% agreement between examiners (p<0.001). Fetal characteristics at preoperative evaluation differed significantly by lesion type, including lesion volume (p<0.001), largest ventricle size (p=0.008), tonsillar herniation (p=0.005), and head circumference (p=0.03). Lesion level, talipes, and lower extremities movement did not differ by type. Surgical and perinatal outcomes differed by lesion type, including need for patch skin closure (p<0.001), gestational age at delivery (p=0.01), and NICU length of stay (p<0.001). PPROM, CSF leakage at birth, and CSF diversion in the NICU did not differ between lesion groups. Linear regression associated severity of ventriculomegaly with lesion type, but not with tonsillar herniation level.

**Conclusions and Relevance:** There is a distinct phenotypic spectrum in open spina bifida with differential baseline presentation and outcomes. Severity of ventriculomegaly is associated with lesion type, rather than tonsillar herniation level. Our findings expand the classification of spina bifida to reveal a spectrum that warrants further study.

**Key Points:** *Question:* Are variations in spina bifida lesion morphology associated with clinical presentationand outcomes?

*Findings:* This secondary analysis of intraoperative images and video from fetoscopic spina bifida repair surgery distinguished five types of spina bifida lesion based on extent of nerve root stretching and neural placode exposure. Preoperative fetal characteristics and postoperative outcomes were significantly associated with lesion type.

*Meaning:* The novel classification of spina bifida to reveal a spectrum with clinical and research implications.

## 4. Introduction

Spina bifida, the most common open neural tube defect in humans compatible with life, develops when a fetus’s neural tube fails to close within the first four weeks after conception.^1,2^ The resulting lesion is conventionally typed as myelomeningocele or myeloschisis, based on the presence or absence, respectively, of a cerebrospinal fluid (CSF)–filled sac over the open neural tube.^3^ In myelomeningocele, this sac protects the covered portion of the spinal cord from exposure to amniotic fluid but stretches the nerve roots. Both lesion types are characterized by CSF leakage, and the developing central nervous system’s exposure to amniotic fluid results in hydrocephalus, hindbrain herniation (Chiari type II malformation), and consequent long-term morbidity and mortality.^4^ These two lesion types represent a spectrum of morphologies, however, as neural placode exposure to amniotic fluid in utero and nerve root stretching at the lesion site vary by degree.^3^ Clinical features also vary, including severity of ventriculomegaly and loss of lower extremities function.

Spina bifida’s formation and effects have been explained by a two-hit hypothesis, wherein the initial defect that prevents neural tube closure is followed by chemical or mechanical damage to the exposed neural placode in utero.^5^ Reducing the placode’s exposure to amniotic fluid to minimize intrauterine damage and reverse hindbrain herniation provided the rationale for repairing spina bifida prenatally.^6^ The randomized Management of Myelomeningocele Study (MOMS) showed that patients who underwent prenatal surgery to repair spina bifida needed ventriculoperitoneal shunting less often and had better spinal cord function than those who underwent postnatal repair.^7^ Since MOMS, prenatal spina bifida repair via open hysterotomy has become a widely accepted treatment option, and less invasive, fetoscopic repair techniques have been tested with promising results.^8,9^ Nonetheless, children who undergo in-utero spina bifida repair do not all benefit: in the MOMS cohort, 58% could not ambulate independently at 30 months, and 68% needed shunt placement.^7^

Understanding the factors that contribute to this variability in long-term benefits after inutero spina bifida repair is essential to improving clinical outcomes. Although recent retrospective studies have shown that fetal and neonatal motor function vary according to spina bifida lesion type,^3,10,11^ a detailed typology of spina bifida lesions, characteristics and associated outcomes is currently lacking.

In-utero repair provides an opportunity to directly visualize and assess spina bifida lesion morphology midway through gestation. This study leveraged this opportunity to identify and characterize a spectrum of spina bifida lesion types and correlate them with preoperative presentation and postoperative outcomes.

## 5. Methods

### Study Design

We conducted a secondary analysis of intraoperative images and video from fetoscopic spina bifida repair surgery to identify lesion types based on morphology and correlate these types with preoperative characteristics and postoperative outcomes.

### Study Population

A prospective cohort of patients underwent fetoscopic spina bifida repair under approved protocols between 2020 and 2023. Patients referred to the UTHealth Houston Fetal Center for suspected fetal spina bifida underwent a comprehensive ultrasound evaluation, fetal magnetic resonance imaging (fMRI), and multidisciplinary counseling. Fetoscopic in-utero spina bifida repair was offered to those who met MOMS trial inclusion criteria,^7^ with maternal body mass index (BMI) limit extended from 35 kg/m^2^ to 45 kg/m^2^ and amniotic fluid alpha fetoprotein higher than 2.5 multiples of the median and positive for acetylcholinesterase to confirm CSF leakage into the amniotic fluid. Eligible patients were offered participation in an institutional review board (IRB)-approved spina bifida registry and an FDA- and IRB-approved trial studying the feasibility of laparotomy-assisted fetoscopic spina bifida repair using a cryopreserved human umbilical cord allograft as a meningeal patch (NCT06042140). Patients were counseled about risks, benefits and alternatives, and informed consent was obtained before surgery. High-resolution images and videos were collected during surgery per protocol. Patients with no images available were excluded from this analysis.

### Preoperative evaluation

Patients underwent ultrasound examinations by certified sonographers utilizing GE E8 and E10 (GE Voluson Expert Ultrasound Equipment; GE Healthcare Ultrasound, Milwaukee, WI) systems. Lesions were measured in three dimensions: anterior-posterior (posterior vertebral body surface to the sac surface), transverse (widest dimension of the sac) and vertical (cephalo-caudad). In lesions with no sac, the vertical and transverse dimensions were measured from the skin defect. The anterior-posterior dimension was measured as zero if there was no displacement of the sac. Fetal lower extremity movements were evaluated, with normal movements defined as any movement at the hips, knees, or ankles over 30 minutes. Right and left lateral ventricles were measured in the posterior horn at the choroid plexus. Tonsillar herniation was evaluated by fMRI per established methods.^12,13^

### Fetoscopic spina bifida repair

In-utero spina bifida repair was performed via laparotomy-assisted fetoscopic approach, using cryopreserved human umbilical cord allografts (HUC-NEOX Cord 1K^®^, Tissue Tech Inc, Miami, FL) as a meningeal patch over the spinal placode, followed by skin closure (eMethods). ^14^ If the skin defect was too large to approximate the edges together, then a second HUC patch was used for skin closure. Video and images were captured via high-resolution Hopkins^®^ 45° Telescope 5 mm (Karl Storz Endoscopy America, Inc.) used for visualization.

After surgery, patients recovered in hospital for 2-3 days. Patients stayed in Houston for management at The Fetal Center or returned home for delivery. Neonatal care was managed according to standard care.

### Outcomes

Duration of surgery and need for patch skin closure were recorded. Perinatal outcomes included gestational age at delivery, preterm premature rupture of membranes (PPROM), and CSF diversion in the neonatal intensive care unit (NICU) (at the pediatric neurosurgeon’s discretion). For patients who delivered outside The Fetal Center, perinatal outcomes were collected by delivery record review and calls to patients.

### Lesion classification

By reviewing intraoperative images and videos, we distinguished five lesion types based on nerve root stretching and placode exposure (Figure 1).

- **Type 1:** Large sac, neural placode displaced from the spinal canal and attached to the highest portion of the protruded thecal sac. Central canal is open through a small opening with skin and meningeal coverage of the sac. Minimal neural placode exposure to amniotic fluid. Nerve roots are stretched from the spinal canal.
- **Type 2:** Large sac, the placode displaced from the spinal canal to the highest portion of the protruded thecal sac. The placode is open for more than half the lesion’s vertical length, with direct exposure to amniotic fluid. Nerve roots are stretched from the spinal canal.
- **Type 3:** Like Type 2, but less placode displacement and less nerve root stretching. The placode is open less than half the lesion’s vertical length, with less exposure to amniotic fluid.
- **Type 4:** The placode is not displaced from the vertebral canal, and nerve root stretching is minimal. The placode is open, but some meningeal coverage reduces exposure to amniotic fluid.
- **Type 5:** Like Type 4, but no meningeal coverage. The placode is completely exposed to amniotic fluid. Nerve roots are not stretched from the spinal canal.

**Figure 1.**
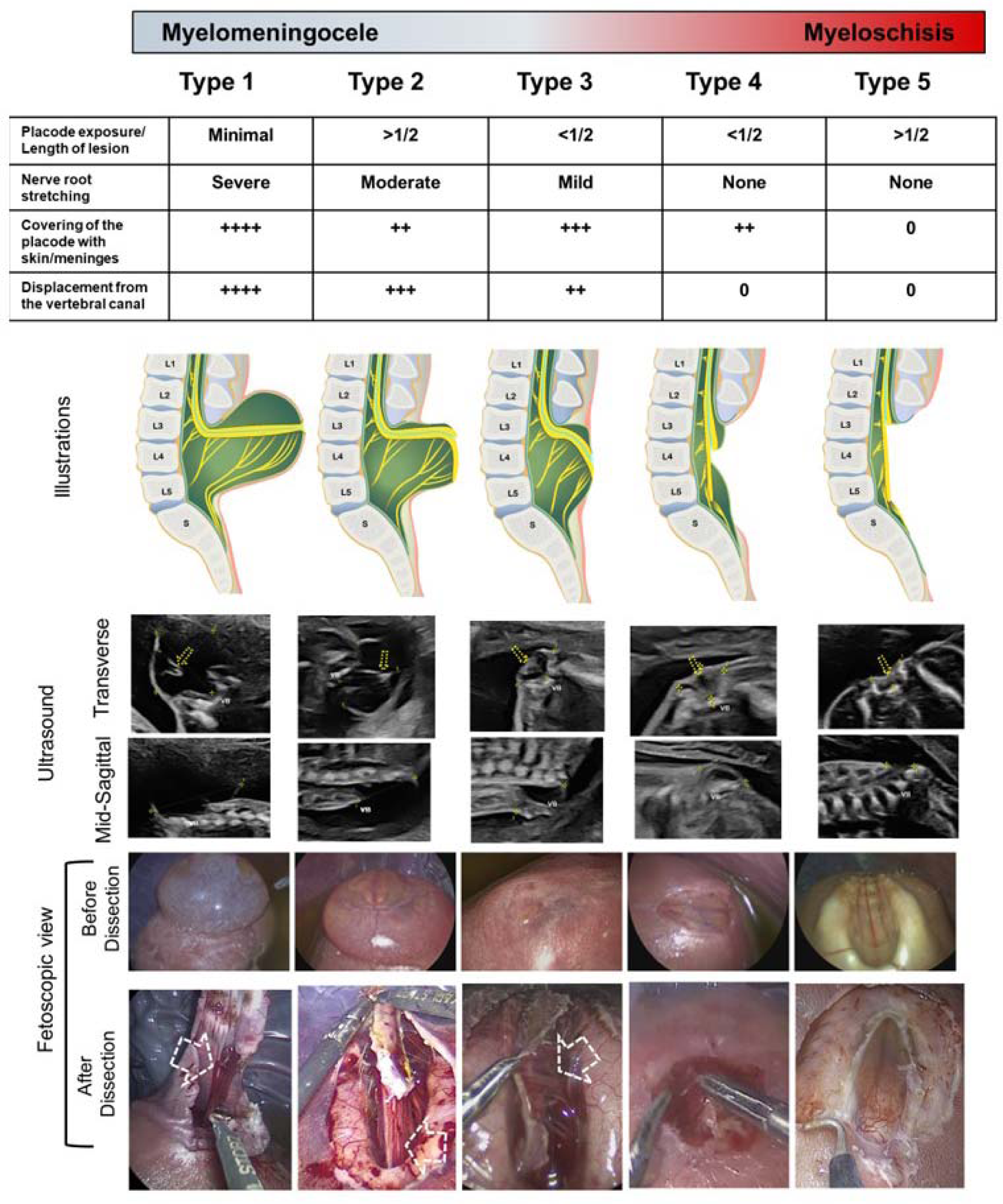
Morphometric classification of spina bifida lesions. Using high-resolution fetoscopic images and videos obtained during surgery, we distinguished five lesion types based on stretching of the nerve roots and exposure of the neural placode. The table presents the key findings used to distinguish the category based on the visual characteristics observed in the fetoscopic view. The illustrations show the 5 types, demonstrating the differences in nerve root stretching and placode exposure to amniotic fluid (– *Blue line*: dura mater; – *Green* line: arachnoid layer; – *Black line*: pia mater; – *Pink line*: skin layer; *Yellow color:* Spinal cord and nerve roots; L: lumbar vertebra; S: sacral vertebra). Two rows of ultrasound images present (Top) transverse images at the mid-lesion and (Bottom) midline sagittal images of the lesion by lesion type. Yellow dotted arrows indicate the neural placode with dorsal displacement/ectopia from the vertebral canal. Two rows of fetoscopic images present lesions (Top) before and (Bottom) after dissection by lesion type. White dotted arrows indicate stretching nerve roots.

The primary reviewer (RP) classified each lesion retrospectively based on these criteria. Then, a second reviewer (LKM) independently classified each lesion by the same criteria for reproducibility. Both were blinded to preoperative and postoperative findings and outcomes. Disagreement cases are presented in the Results.

### Statistical Analysis

Data were analyzed and visualized in R using ggpubr and ggplot2 packages. Descriptive, then inferential statistics were reported. We derived the semi-ellipsoid lesion volume by the equation, 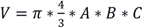, where A, B, and C are lesion dimensions. Due to non-normally distributed data, the Kruskal-Wallis test was performed. Mann–Whitney U test for multiple comparisons was used for post hoc analysis to determine significant main effects. Categorical data were compared via Fisher’s Exact test. Interrater agreement for lesion types was assessed using kappa statistics. Preoperative ventricle size and its association with lesion type, tonsillar herniation and lesion level were evaluated via linear regression in SAS (9.4, Cary, NC).

## 6. Results

Of 60 spina bifida lesions repaired fetoscopically at The Fetal Center from 2020–2023, 57 had high-quality images available. Forty-eight patients were enrolled through an FDA feasibility study; nine were enrolled as practice of medicine cases in an observational cohort study. Nineteen patients delivered outside The Fetal Center.

### Lesion classification

Lesion types 1 through 5 accounted for 14%, 39%, 16%, 14%, and 18%, respectively, of the lesions in our cohort (Table 1). Reviewers classified lesions with 93% agreement (p<0.001). They disagreed in three cases: Type 4 and Type 5; Type 2 and Type 3; Type 2 and Type 5. The third disagreement case was reviewed: the second reviewer’s interpretation was affected by the angle of the lesion image. The primary reviewer’s classification was selected for the final analysis in all cases.

**Table 1.**
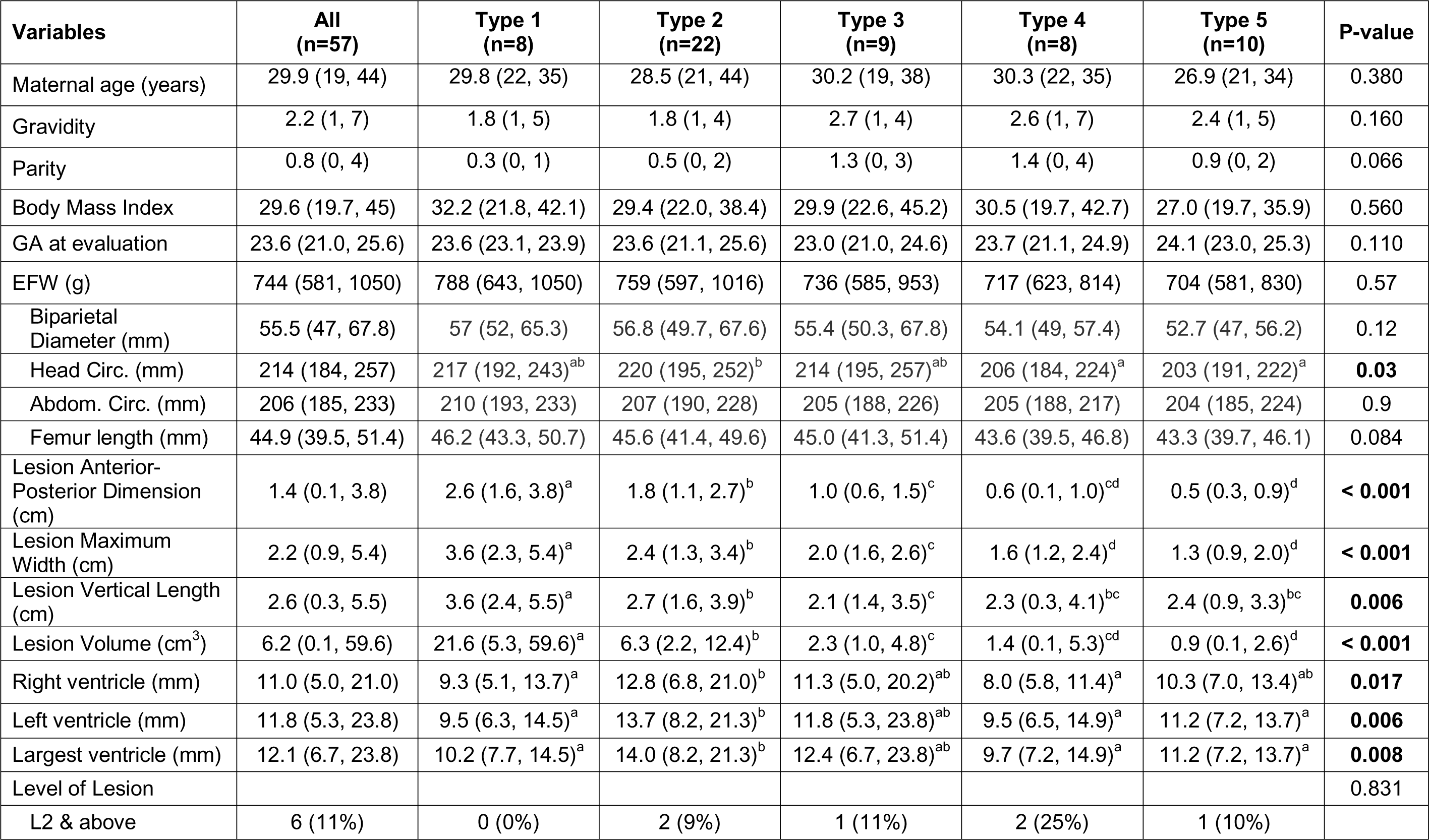
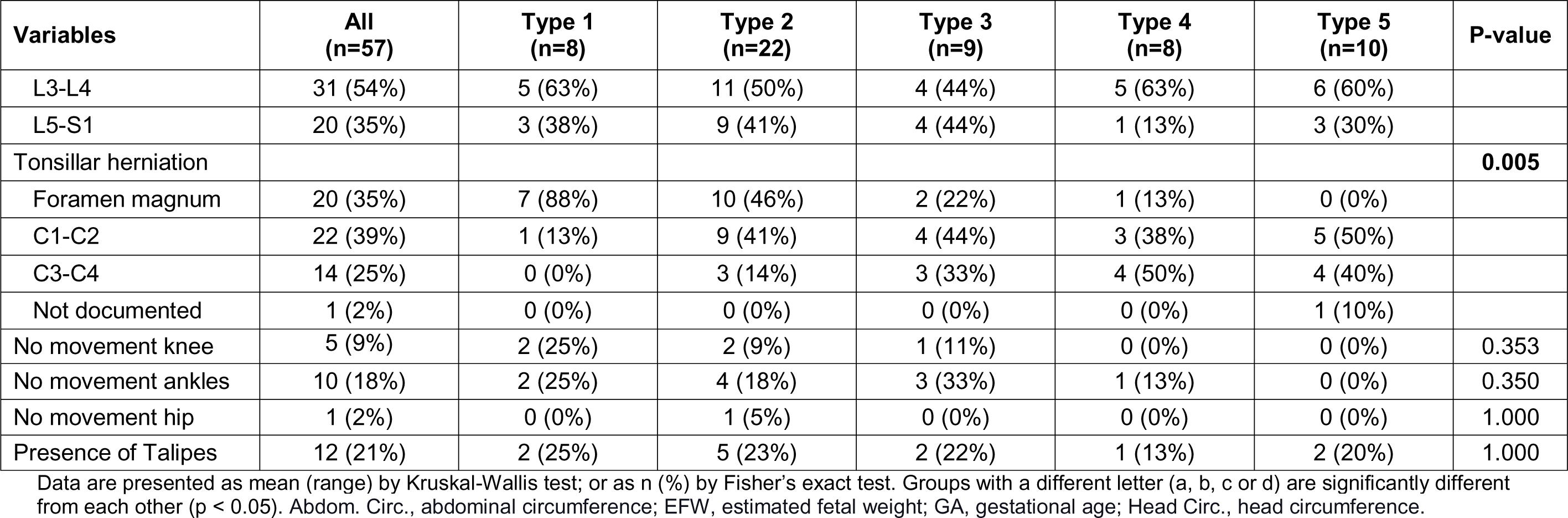
Maternal demographics and preoperative characteristics by lesion type.

### Preoperative characteristics

Maternal demographics did not differ between lesion groups, nor did fetal extremity movement, lesion level, or talipes (Table 1). Estimated fetal weight (EFW) did not differ by lesion type when considered as an aggregate measure, but analysis of EFW’s subcomponents found significant differences in head circumference (HC) by lesion type (p=0.03): fetuses with Type 2 lesions had a higher percentile HC than those with Types 4 (p<0.05) and 5 (p<0.001), and those with Types 1 and 3 had a higher percentile HC than those with Type 5 (p<0.01) (Figure 2).

**Figure 2.**
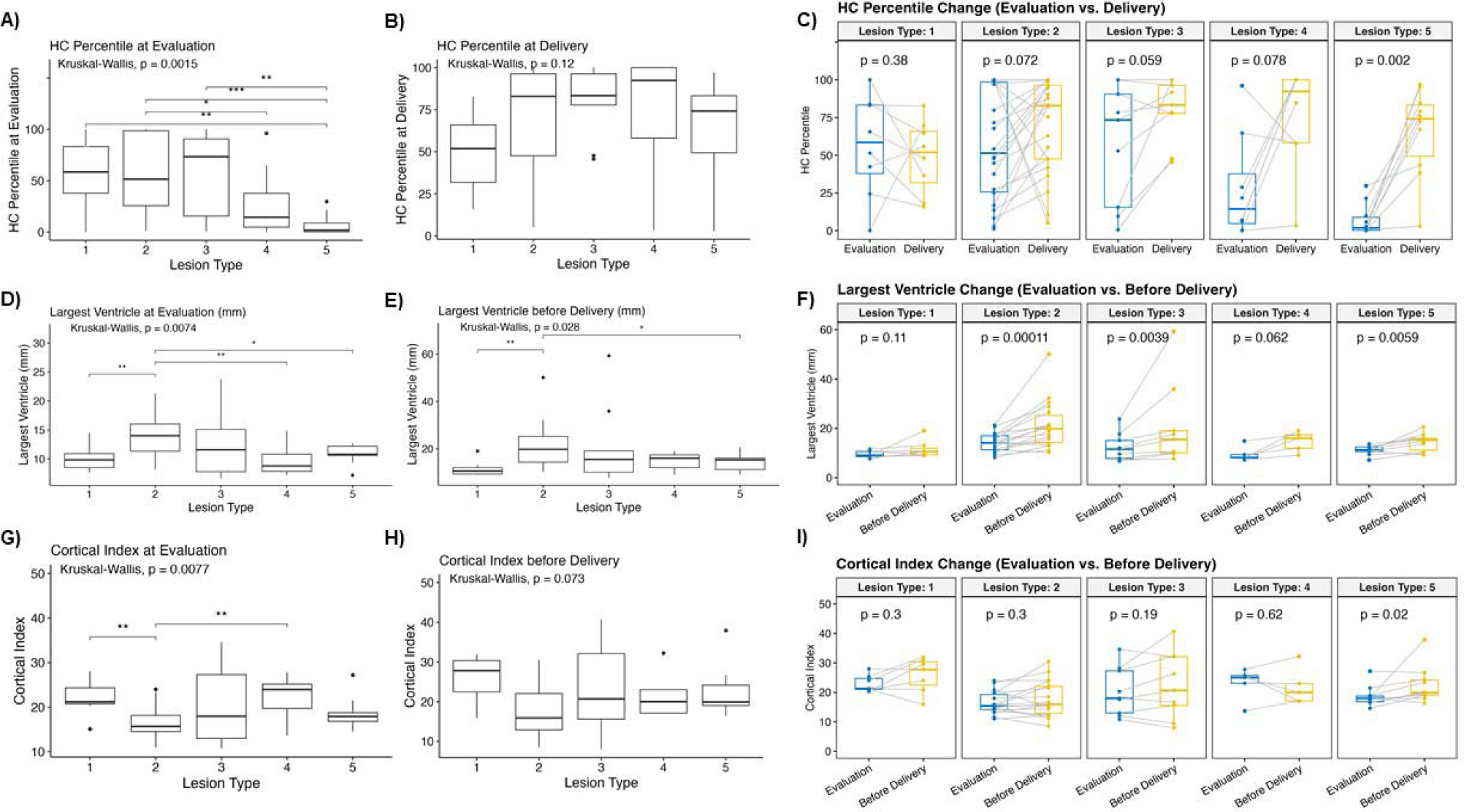
Fetal head biometry differs by lesion type at baseline evaluation and changes after fetal surgery. Box plots compare fetal **(A-C)** head circumference (HC) percentile, **(D-F)** largest ventricle size, and **(G-I)** Cortical Index (CI) by lesion type at preoperative evaluation **(A, D, G)** and at delivery **(B, E, H)** and show the change between timepoints **(C, F, I)**. CI was calculated as HC (in mm) divided by ventricle size (in mm). Within each box, horizontal bold black lines denote median values; boxes extend from the 25th to the 75th percentile of each group’s distribution of values; vertical extending lines denote adjacent values (i.e., the most extreme values within 1.5 interquartile range of the 25th and 75th percentile of each group); dots denote observations outside the range of adjacent values. *p<0.05, **p<0.01.

Lesion dimensions differed by type (Table 1; eFigure 1). Type 1 lesions were significantly larger than all other types in all three dimensions and in volume. Type 2 lesions were longer than Type 3 vertically and larger than Types 3-5 in the anterior-posterior and transverse dimensions and in volume. Type 3 was wider than Type 4 and larger than Type 5 in the anterior-posterior and transverse dimensions and in volume.

Largest ventricle size differed by lesion type (p=0.0074), as fetuses with Type 2 lesions had larger ventricles than those with Types 1, 4, and 5 (Table 1, Figure 2). Right (p=0.017) and left (p=0.0058) ventricle sizes differed similarly by lesion type: Type 2 had larger right ventricles than Types 1 and 4 and larger left ventricles than Types 1, 4, and 5 (Table 1; eFigure 2A, 2C). When adjusting for HC, Type 2 had larger right and left ventricles than Types 1 and 4; Type 5 had larger right ventricles than Type 4 and larger left ventricles than Types 1 and 4 (eFigure 2B, 2D).

### Ventricle size is associated with lesion type

We evaluated the association between ventricle size and lesion type, lesion level, and tonsillar herniation via linear regression. Conditioning on the lesion level and tonsillar herniation (foramen magnum or above: mild; C1-C2: moderate; C3-C4: severe), largest ventricle size is significantly associated with lesion type (Table 2). Specifically, largest ventricle is greater in Type 2 than in Type 5 lesions (p=0.008). Fetuses with lower lesion levels had smaller ventricles, but this was not statistically significant (p=0.07).

**Table 2.** Linear regression analysis of factors associated with preoperative ventricle size.

| **Independent Variables** | **Beta** | **Standard Error** | **t-Value** | **P-Value** |
| --- | --- | --- | --- | --- |
| Lesion Type 1 | 0.664 | 1.877 | 0.35 | 0.7252 |
| Lesion Type 2 | 3.959 | 1.432 | 2.76 | **0.0080** |
| Lesion Type 3 | 1.960 | 1.599 | 1.23 | 0.2261 |
| Lesion Type 4 | -1.641 | 1.652 | -0.99 | 0.3253 |
| Lesion Level | -1.371 | 0.743 | -1.84 | 0.0712 |
| Tonsillar Herniation | 0.896 | 0.717 | 1.25 | 0.2174 |

All lesion types are compared with lesion Type 5.

### Surgical and neonatal outcomes

All 10 (100.0%) Type 5 lesions required skin patch closure, which differed from all other lesion types (p<0.001). Surgical time and gestational age at surgery did not differ between groups, nor did PPROM, CSF leakage at birth, wound revision, or CSF diversion in the NICU (Table 3).

**Table 3.**
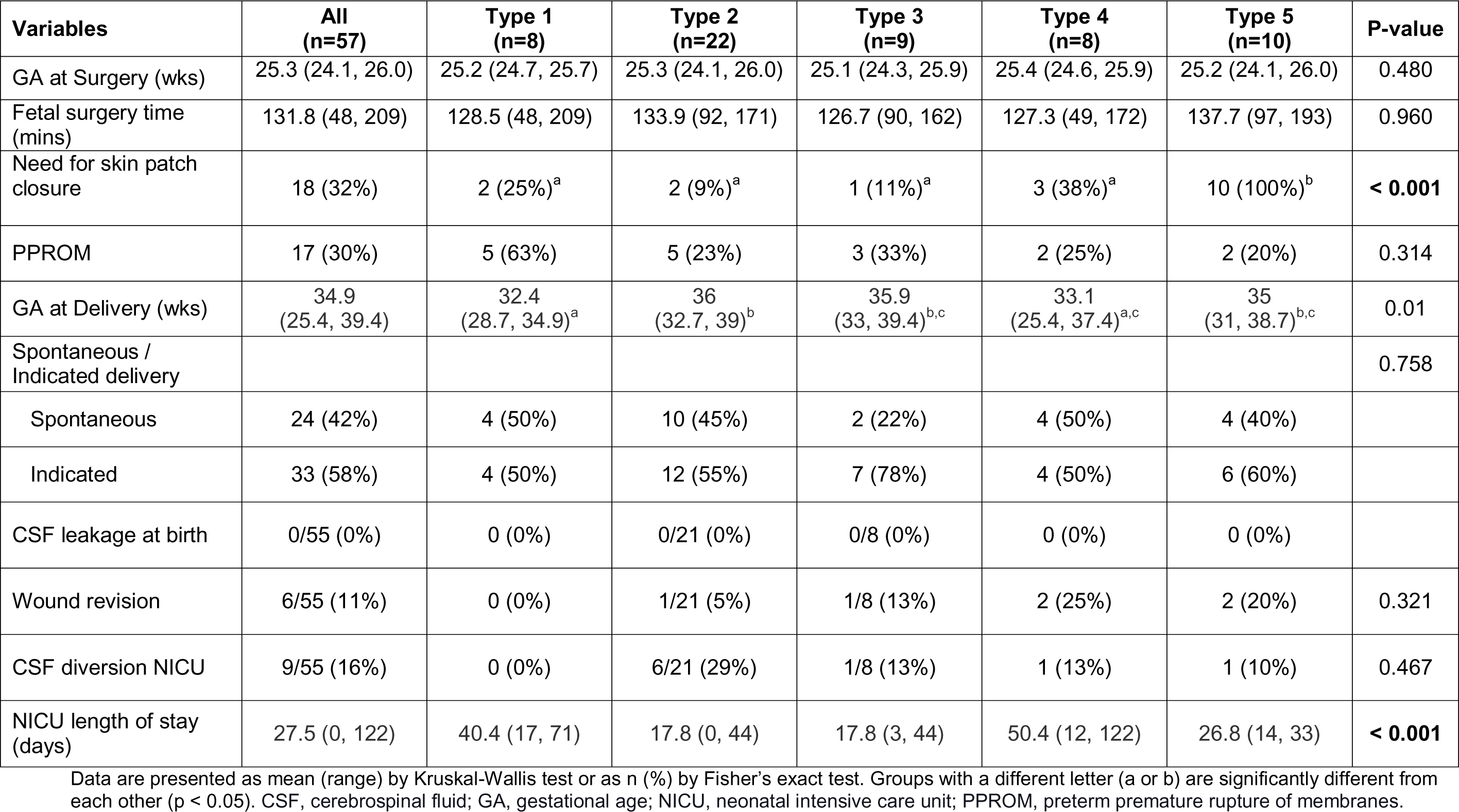
Postoperative and neonatal outcomes by lesion type.

| Variables | All<br>(n=57) | Type 1<br>(n=8) | Type 2<br>(n=22) | Type 3<br>(n=9) | Type 4<br>(n=8) | Type 5<br>(n=10) | P-value |
| --- | --- | --- | --- | --- | --- | --- | --- |
| GA at Surgery (wks) | 25.3 (24.1, 26.0) | 25.2 (24.7, 25.7) | 25.3 (24.1, 26.0) | 25.1 (24.3, 25.9) | 25.4 (24.6, 25.9) | 25.2 (24.1, 26.0) | 0.480 |
| Fetal surgery time<br>(mins) | 131.8 (48, 209) | 128.5 (48, 209) | 133.9 (92, 171) | 126.7 (90, 162) | 127.3 (49, 172) | 137.7 (97, 193) | 0.960 |
| Need for skin patch<br>closure | 18 (32%) | 2 (25%)<sup>a</sup> | 2 (9%)<sup>a</sup> | 1 (11%)<sup>a</sup> | 3 (38%)<sup>a</sup> | 10 (100%)<sup>b</sup> | **< 0.001** |
| PPROM | 17 (30%) | 5 (63%) | 5 (23%) | 3 (33%) | 2 (25%) | 2 (20%) | 0.314 |
| GA at Delivery (wks) | 34.9<br>(25.4, 39.4) | 32.4<br>(28.7, 34.9)<sup>a</sup> | 36<br>(32.7, 39)<sup>b</sup> | 35.9<br>(33, 39.4)<sup>b,c</sup> | 33.1<br>(25.4, 37.4)<sup>a,c</sup> | 35<br>(31, 38.7)<sup>b,c</sup> | 0.01 |
| Spontaneous /<br>Indicated delivery |  |  |  |  |  |  | 0.758 |
| Spontaneous | 24 (42%) | 4 (50%) | 10 (45%) | 2 (22%) | 4 (50%) | 4 (40%) |  |
| Indicated | 33 (58%) | 4 (50%) | 12 (55%) | 7 (78%) | 4 (50%) | 6 (60%) |  |
| CSF leakage at birth | 0/55 (0%) | 0 (0%) | 0/21 (0%) | 0/8 (0%) | 0 (0%) | 0 (0%) |  |
| Wound revision | 6/55 (11%) | 0 (0%) | 1/21 (5%) | 1/8 (13%) | 2 (25%) | 2 (20%) | 0.321 |
| CSF diversion NICU | 9/55 (16%) | 0 (0%) | 6/21 (29%) | 1/8 (13%) | 1 (13%) | 1 (10%) | 0.467 |
| NICU length of stay<br>(days) | 27.5 (0, 122) | 40.4 (17, 71) | 17.8 (0, 44) | 17.8 (3, 44) | 50.4 (12, 122) | 26.8 (14, 33) | **< 0.001** |

Neonates with Type 1 lesions were delivered at an earlier gestational age than those with Types 2, 3, and 5; those with Type 4 lesions were delivered earlier than those with Type 2 (Table 3, eFigure 3A). Neonates with Type 2 and 3 lesions required fewer days in the NICU than those with Types 1 and 4; Type 2 also had fewer NICU days than Type 5 (p<0.001) (eFigure 3B).

### Cortical Index from baseline to delivery

Despite significant differences in HC percentile at baseline among lesion groups (p=0.0015), HC percentile at delivery did not differ by lesion type (p=0.12) (Figure 2). By contrast, largest ventricle size at last ultrasound (within four weeks of delivery; n=49) differed by lesion type (p=0.028): Type 2 had larger ventricles than Types 1 and 5. Analysis of the change in HC percentile from baseline to delivery found a marked increase among those with Type 5 lesions (p=0.002). Types 2–4 increased in HC, but not statistically significantly. Ventricle size increased in Types 2 (p=0.00011), 3 (p=0.0039), and 5 (p=0.0059).

To determine the extent to which the increase in HC from baseline to delivery reflects cortical development versus ventricle growth, we calculated the Cortical Index (CI) by dividing HC (in mm) by ventricle diameter (in mm).^15^ CI at baseline differed significantly by lesion type (p=0.0077): Type 2 had a lower CI than Types 1 and 4, consistent with ventricular differences. CI before delivery did not differ by lesion type (p=0.073). CI increased from baseline in Type 5 (p=0.02), but not in other lesion groups (Figure 2).

## 7. Discussion

Spina bifida is conventionally classified as myelomeningocele or myeloschisis, based on the presence or absence, respectively, of a sac covering the open neural tube.^10^ However, our clinical observations from performing spina bifida repair at a high-volume center suggested a range of lesion morphologies inconsistent with a binary classification. Clinical features, fetal anatomy, and postnatal outcomes also vary considerably. Understanding the factors that contribute to this variability requires a more complex classification of spina bifida lesion types and their association with preoperative presentation and postoperative outcomes. As a step toward that classification, we evaluated intraoperative images and videos, and we distinguished five types of spina bifida lesions based on nerve root stretching and neural placode exposure. Fetal characteristics at baseline evaluation, including head circumference and ventricle size, differed significantly by these lesion types. Operative and perinatal outcomes, including need for patch skin closure, gestational age at delivery, and length of NICU stay, also differed by lesion type. Collectively, these findings suggest a spectrum of spina bifida types that warrants further characterization. These findings are potentially significant for clinical practice, as understanding the range of spina bifida presentations and their associated outcomes would inform patient counseling and surgical planning.

In the study, lesion type was the most important association factor for ventriculomegaly, with Type 2 having the largest ventricular diameter. However, there was no association between ventricle size and tonsillar herniation level. This differs from the common understanding that hindbrain herniation obstructing cerebrospinal fluid flow through the foramen Luschka and Magendie in the fourth ventricle contributes to ventriculomegaly.^19^ Future studies in a larger cohort would allow group analyses of the relation between ventricle size, lesion level, and hindbrain herniation within each lesion type.

Our analysis of fetal characteristics and perinatal outcomes by lesion type revealed significant gains in head circumference (HC), from around the 5^th^ to the 75^th^ percentile, among those with Type 5 lesions. That group’s corresponding increase in Cortical Index (CI), reflecting faster growth of HC than of ventricles, suggests that the changes in HC are not attributable to ventriculomegaly alone and may indicate cortical mass development after in-utero spina bifida repair. Similarly, Danzer et al. reported fetal head biometry changes with increases in CI after in-utero repair in 50 fetuses.^15^ In our cohort, fetuses with lesion Types 2-4 also showed increases in HC from baseline to delivery, but these were not statistically significant, nor did CI change significantly in those groups. One possible explanation is that fetuses with Type 2-4 lesions may have narrowing of the cerebral aqueduct that does not improve with in-utero spina bifida repair. Other recent analyses have reported differences in motor function between different lesion types. Oliver et al.’s review of 404 patients found that myelomeningocele lesions, whether repaired prenatally or postnatally, are associated with fetal talipes and impaired lower extremities movement.^3^ Corroenne et al. reported that fetuses with myeloschisis were 3.1 times more likely to have intact motor function prenatally^10^ and 11.2 times more likely to have intact motor function at birth.^11^ Farmer et al.’s study of pediatric outcomes in the MOMS cohort reported that absence of a sac covering the lesion was associated with independent ambulation at 30 months among patients who underwent prenatal surgery.^18^ The extent to which differences in presentation and outcomes across lesion types reflect anatomical variations versus response to therapy or other factors requires further investigation. The lack of significant findings in several of the lesion types could be due to the smaller sample sizes in those types. Larger studies are needed to further explain these findings.

Our demonstration of a broader spectrum of lesion types than previously assumed may have implications for future molecular studies, allowing insight into genetic and epigenetic mechanisms contributing to lesion development. Spina bifida is believed to be highly heritable, but its complex etiology, rarity in the population and phenotypic heterogeneity make variant detection and gene/pathway discovery challenging. Studies have identified deleterious genomic variants that may increase the risk for developing spina bifida.^20,21^ Others have demonstrated the importance of studying the genomic and transcriptomic profiles of spina bifida lesions.^22–24^ Nonetheless, further studies are needed to explore the molecular basis for variations in spina bifida lesion formation and its clinical implications.

Our study’s major strength is its prospective, consistent data collection under an FDA clinical trial protocol. All surgeries were performed at a single, high-volume center, thus ensuring procedural consistency. Our study population was recruited under broad inclusion criteria comparable to the MOMS trial. We confirmed the interrater reliability of our classification between two independent reviewers. This study was limited by a small sample size not powered to compare differences between groups that were not significant, such as lower extremities movement. The lesion types of those not offered prenatal surgery may differ from those who underwent surgery. Differences in neonatal management across centers may have affected outcomes for patients who delivered outside The Fetal Center. There was no identifiable reason for preterm birth in the Type 1 group. This needs further evaluation in a larger cohort.

## Conclusions

Direct visualization of spina bifida lesions during fetoscopic repair surgery distinguished five types of spina bifida that differ with respect to nerve root stretching and neural placode exposure to amniotic fluid. These types may have implications for baseline function and prognosis. Further research is needed to explore the molecular basis for variations in spina bifida formation and response to in-utero therapy.

## Data Availability

All data produced in the present study are available upon reasonable request to the authors

## 8. Acknowledgements

Lovepreet K. Mann and Ramesha Papanna had full access to all the data in the study and take responsibility for the integrity of the data and the accuracy of the data analysis. The authors would like to acknowledge Arthur Day, MD, Bradley E. Weprin, MD and John Honeycutt, MD for their assessments, as independent pediatric neurosurgeons, of the completeness of the surgical closure for the FDA IDE study.

