## Supplement for "Morphometric Analysis of Spina Bifida after Fetal Repair Shows New Subtypes with Associated Outcomes"

**Supplementary Online Content**

**eMethods.** Fetoscopic Spina Bifida Repair Procedures.

**eFigure 1.** Lesion dimensions differ by lesion type.

**eFigure 2.** Baseline ventricle size differs by lesion type.

**eFigure 3.** Gestational age at delivery and length of NICU stay differ by lesion type.

This supplementary material has been provided by the authors to give readers additional information about their work.

**eMethods. Fetoscopic spina bifida repair procedures.**

In-utero spina bifida repair was performed via a laparotomy-assisted fetoscopic approach. Before surgery, patients received tocolysis via magnesium drip dispensing a 6 g bolus over 30 min, followed by a continuous infusion of 2 g/hr throughout the operation. An epidural catheter was placed for postoperative pain management, and maternal general anesthesia was administered via endotracheal intubation with sevoflurane, propofol, fentanyl and rocuronium.

The uterus was exteriorized through a vertical lower midline abdominal skin incision, then the fetus was positioned by external cephalic version. The placental location was then mapped via ultrasound, and three ports (9 Fr, 16 Fr and 16 Fr or 5 mm) were placed in the uterine cavity away from the placenta. The amniotic fluid was partially removed, and the uterus was insufflated with heated-humidified carbon dioxide (hhCO_2_) with a pressure of 8-12 mmHg.

Fetal anesthesia was administered in the fetal presenting thigh as an intramuscular injection of fentanyl (20 mcg/kg) and vecuronium (0.2 mg/kg) every 90 minutes. Lesion images were taken prior to placode dissection. Then, the placode was dissected sharply with laparoscopic microscissors and microtip bovie, circumferentially around the placode without damaging the spinal cord. The junctional zone between the meninges and the skin was circumferentially excised. The spinal cord placode was then covered with cryopreserved human umbilical cord allografts (HUC-NEOX Cord 1K^®^, Tissue Tech Inc, Miami, FL) and sutured to the underlying muscle and fascia circumferentially using 3-0 self-anchoring bidirectional suture (Stratafix^TM^, Ethicon, Johnson & Johnson, New Brunswick, NJ) followed by primary closure of the skin. If the skin defect was large and the edges could not be approximated together, then a second human umbilical cord patch was used for skin closure and circumferentially sutured with absorbable suture. At the end of the procedure, the hhCO2 gas was removed, and 1 g of Vancomycin in warm lactated Ringer’s solution was replaced into the uterine cavity. The uterine wall ports were closed using anchoring sutures. The abdominal wall was closed in layers.

**eFigure 1. Lesion dimensions differ by lesion type.**

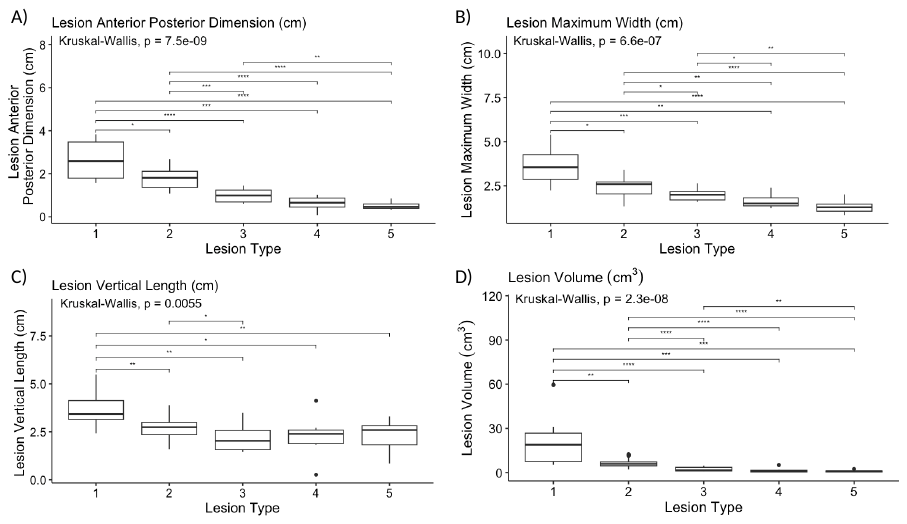

Box plots compare lesions’ **A)** anterior-posterior dimension, **B)** maximum width, **C)** vertical length, and **D)** volume by lesion type. Within each box, horizontal bold black lines denote median values; boxes extend from the 25th to the 75th percentile of each group's distribution of values; vertical extending lines denote adjacent values (i.e., the most extreme values within 1.5 interquartile range of the 25th and 75th percentile of each group); dots denote observations outside the range of adjacent values. *p<0.05, **p<0.01, ***p<0.001, ****p<0.0001.

**eFigure 2. Baseline ventricle size differs by lesion type.**

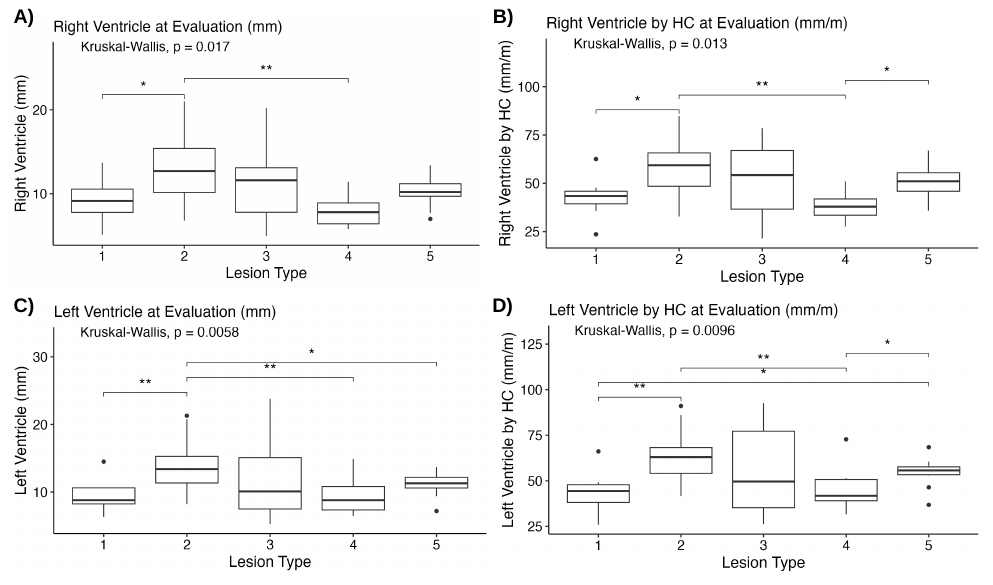

Box plots compare **A)** right ventricle size, **B)** right ventricle size adjusted for head circumference, **C)** left ventricle size, and **D)** left ventricle size adjusted for head circumference by lesion type. Within each box, horizontal bold black lines denote median values; boxes extend from the 25th to the 75th percentile of each group's distribution of values; vertical extending lines denote adjacent values (i.e., the most extreme values within 1.5 interquartile range of the 25th and 75th percentile of each group); dots denote observations outside the range of adjacent values. *p<0.05, **p<0.01.

**eFigure 3. Gestational age at delivery and length of NICU stay differ by lesion type.**

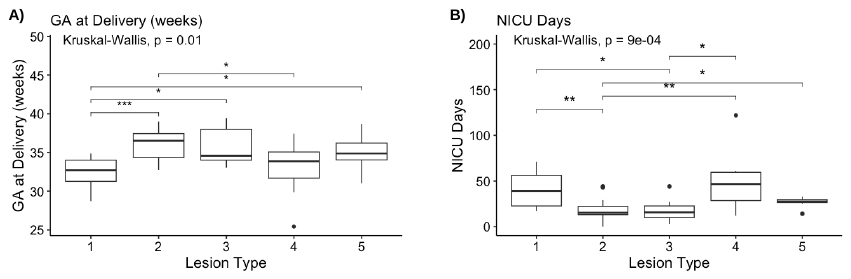

Box plots compare **A)** gestational age (GA) at delivery and **B)** days in the neonatal intensive care unit (NICU) by lesion type. Within each box, horizontal bold black lines denote median values; boxes extend from the 25th to the 75th percentile of each group's distribution of values; vertical extending lines denote adjacent values (i.e., the most extreme values within 1.5 interquartile range of the 25th and 75th percentile of each group); dots denote observations outside the range of adjacent values. *p<0.05, **p<0.01, ***p<0.001.
